# Contact patterns of older adults with and without frailty in the Netherlands during the COVID-19 pandemic

**DOI:** 10.1101/2023.05.09.23289550

**Authors:** Jantien A Backer, Jan van de Kassteele, Fatima El Fakiri, Niel Hens, Jacco Wallinga

## Abstract

**Background:** During the COVID-19 pandemic, social distancing measures were imposed to protect the population from exposure, especially older adults and people with frailty, who have the highest risk for severe outcomes. These restrictions greatly reduced contacts in the general population, but little was known about behaviour changes among older adults and people with frailty themselves. Our aim was to quantify how COVID-19 measures affected the contact behaviour of older adults and how this differed between older adults with and without frailty.

**Methods:** In 2021, a contact survey was carried out among people aged 70 years and older in the Netherlands. A random sample of persons per age group (70–74, 75–79, 80–84, 85–89, and 90+) and gender was invited to participate, either during a period with stringent (April 2021) or moderate (October 2021) measures. Participants provided general information on themselves, including their frailty, and they reported characteristics of all persons with whom they had face-to-face contact on a given day over the course of a full week.

**Results:** In total, 720 community-dwelling older adults were included (overall response rate of 15%), who reported 16,505 contacts. During the survey period with moderate measures, participants without frailty had significantly more contacts outside their household than participants with frailty. Especially for females, frailty was a more informative predictor of the number of contacts than age. During the survey period with stringent measures, participants with and without frailty had significantly lower numbers of contacts compared to the survey period with moderate measures. The reduction of the number of contacts was largest for the eldest participants without frailty. As they interact mostly with adults of a similar high age who are likely frail, this reduction of the number of contacts indirectly protects older adults with frailty from SARS-CoV-2 exposure.

**Conclusions:** The results of this study reveal that social distancing measures during the COVID-19 pandemic differentially affected the contact patterns of older adults with and without frailty. The reduction of contacts may have led to the direct protection of older adults in general but also to the indirect protection of older adults with frailty.

## Background

When the SARS-CoV-2 virus emerged at the end of 2019, it spread rapidly around the globe because of its ability to transmit pre-and asymptomatically and because the population was completely susceptible to this new virus. The only control option that could immediately mitigate the growing epidemic was the implementation of non-pharmaceutical interventions, including social distancing measures (e.g., closing schools, suspending leisure activities, and working from home). These social distancing measures aim to reduce exposure by reducing contact rates of for instance face-to-face and physical contacts, as each of these contacts could be an at-risk event for transmission. Contact surveys showed that in the first wave of the COVID-19 pandemic, contact rates decreased by more than half in China [1], the UK [2], the Netherlands [3], and other countries worldwide [4–7]. The reduction of contact rates effectively decreased the daily number of cases [8–10].

Social distancing was especially important to reduce exposure of older adults and persons with frailty, as they have the highest probability of hospitalisation and death after SARS-CoV-2 infection [11–13]. Frailty as a health status is a generic concept that can be defined and measured in many ways [14]. In hospital or care settings, the focus is usually on physical frailty, as expressed, for instance, by the frailty index [15] or the Clinical Frailty Scale [16]. In community settings, frailty is often used in a broader sense that also comprises cognitive and psychosocial dimensions, as expressed, for instance, by the Tilburg Frailty Indicator [17] or the Groningen Frailty Indicator [18]. All frailty instruments indicate that frailty increases with age and is generally higher for females than males of the same age [19]. The variation in frailty among persons of the same age and gender is substantial, especially in community-dwelling older adults [20].

Two studies conducted in the Netherlands in 2006-2007 [21] and 2016-2017 [22] showed that participants aged 70 years and older contact on average 7.2 (6.2 - 8.5) and 7.0 (6.0 - 8.4) different individuals per day (mean and 95% bootstrap CI), respectively. Surveys among older adults during the first wave of the COVID-19 pandemic showed a decrease in in-person contacts [23], associated with a decrease in general well-being [24] and an increase in frailty [25]. Contact surveys in the general population also showed a significant reduction of contact rates in older age groups [1–5]. However, none of these contact surveys distinguished participants by frailty.

The contact behaviour of older adults may well differ by frailty. On the one hand, older adults with frailty might have fewer contacts due to physical inabilities or social isolation. On the other hand, they could have more contacts when they require more medical or home care. In general, the number of contacts per person decreases by age [26,27] and can differ by gender [26], but it is unknown how this is affected by frailty. As contacts can be used as a proxy for at-risk events for transmission [28], quantifying these contacts by frailty may lead to a better understanding of exposure and hence disease burden in older adults with and without frailty.

We conducted a contact survey among persons aged 70 years and older in the Netherlands to quantify the number of contacts per person by frailty status. By conducting the survey during two distinct survey periods with stringent and moderate COVID-19 measures, we aimed to gain insight into how older adults with and without frailty change their contact behaviour when faced with lockdown measures.

## Methods

### Study design

Persons aged 70 years and older were invited to participate in the contact survey. For each survey period, invitees were randomly sampled from the Dutch Personal Records Database [29], yielding two independent cross-sectional study populations. The invitees were randomly sampled per age group (70-74, 75-79, 80-84, 85-89, 90+) and gender. The numbers of invitees were chosen to obtain similar numbers of respondents in each stratum, assuming lower response rates in older age groups. The questionnaires were unmarked to minimise the collection of personal data. As a consequence, it was not possible to send reminders to non-responders to increase response.

Each participant was requested to provide some general information and to fill out a contact diary each day during a full week. The general information consisted of participant age, gender, country of birth, education level, household size, and vaccination status for COVID-19, influenza, and invasive pneumococcal disease (IPD). To determine the frailty status of a participant, the Groningen Frailty Indicator (GFI) [18] was used. This indicator consists of 15 questions on physical, cognitive, social, and psychological conditions. When an answer contributes to frailty, it would score 1, or 0 otherwise. A GFI score of 4 or higher (out of 15) indicates a frail status. The GFI was preferred over other frailty indicators because it contains few questions and is easy to fill out.

The contact diary consisted of a contact page for each day of the week, where a participant could report all persons with whom they had face-to-face contact that day. A contact was defined as a conversation of at least a few sentences and/or a physical contact; contacts via telephone or the internet were explicitly excluded. For each contacted person, the participant reported the gender, age (range), whether this person was a household member, whether the contact was physical, such as shaking hands, whether the contact was protected, for example, by a face mask, whether the contact lasted longer than 15 minutes, whether a minimal distance of 1.5 m was kept, and the location(s). The location options included the home of the participant, the home of the contact person, the work place (also for volunteer work), transit, leisure, shop, outside, and other (to be specified). Also, a name or description of the person could be reported, not only for ease of filling out but also to identify repeated contacts with the same person on different days of the week. Participants could indicate when they did not have any contacts on a given day, which was helpful to distinguish between participants without any contacts and participants that did not fill out the contact diary on that day.

Returned questionnaires that lacked general information and/or any filled-out contact day were not included in further data processing. Questionnaires that were suitable for analysis were entered in a database with internal consistency checks, e.g., whether the participant is at least 70 years old and whether no duplicate participant id’s are entered. After data cleaning, the data was reformatted to the standard format for contact surveys on socialcontactdata.org and published online [30]. A full description of all steps in the study design is provided in the supplement (Suppl. S1). All code for data cleaning and analysis is publicly available [31].

### Survey periods

The study encompassed two survey periods. The first survey period in April 2021 featured stringent COVID-19 control measures: education was online for most students of secondary schools and universities; working from home was required when possible; face masks were mandatory in indoor public spaces; bars and restaurants were closed, as were all other cultural and leisure activities; and an evening curfew was in place. Most older adults had already had the opportunity to be vaccinated, but mass vaccination of the remaining adult population had only just started. In the second survey period in October 2021, most control measures had been lifted. Schools and work places were fully open; face masks were only required on public transport; and social, cultural, and leisure venues were accessible with proof of vaccination or a negative test result. All persons aged 18 years and older were eligible for vaccination, and many had been fully vaccinated.

### Analysis

Response rates were calculated by age group and gender for both survey periods. The study population was described by summarising the number of responses by survey period, age group, gender, country of birth, GFI score, household size, and education level. To assess the representativeness of the study population, we investigated the non-response biases of participants not born in the Netherlands and participants in long-term care facilities, as we expected these groups to have low response rates. To this end, we compared the number of participants not born in the Netherlands and the number of participants in long-term care facilities to their expected values, based on the size of these groups living in the Netherlands in 2021 [32,33]. The observed vaccination coverages were compared to the actual vaccination coverages [34–36]. We evaluated which characteristics of the study population determine frailty (Suppl. S2) and checked whether this agrees with literature.

To focus the further analyses on community-dwelling older adults, participants who live in a long-term care facility were excluded. The contact behaviour was described by how many people a participant contacted and in which age classes. For the analysis, only community contacts were included, i.e., contacts with non-household members, as these are the contacts that are mostly affected by control measures. The community contacts were summed over the full week to eliminate any day-of-the-week effect. Participants with fewer than 5 completed days were excluded from the contact analysis. The missing data of participants with 5 or 6 days completed were imputed while taking the effects of the day of the week and fatigue into account (Suppl. S3).

The weekly total of community contacts was assumed to follow a negative binomial distribution, where the log of the mean was modelled with full interactions between frailty (0 or 1), participant age (numeric), participant gender (male or female), and survey period (1 or 2). The COVID-19 vaccination status could also affect the number of community contacts, but this covariate was omitted because the data contains few unvaccinated participants. We assessed whether household size (1 or 2+) should have been included as an explanatory variable by comparing the likelihoods of the models with and without household size using a likelihood ratio chi-square test.

In a similar way, the effect of frailty in a specific period and the effect of the period on a specific frailty status were determined using subsets of the data. For example, the effect of frailty in the first survey period was assessed by comparing the likelihood of the model without frailty as an explanatory variable to the likelihood of the model with frailty, using only the data collected in the first survey period. If the likelihoods were similar (according to the chi-square test), the number of contacts did not significantly differ between people with and without frailty in the first survey period. The expected weekly total of community contacts was compared for an average person with and without frailty in the Dutch 70+ population by survey period.

To study age-specific mixing patterns, contact matrices were constructed by dividing the total number of contacts per week between two age groups by the number of participants in the participant age group. The resulting matrix of average numbers of contacts per week per participant is asymmetrical due to the skewed population distribution. We assumed people with and without frailty mix proportionally and divided the matrix by the Dutch population distribution in 2021 [37]. Assuming reciprocal contacts, diagonally opposed matrix elements were averaged, yielding a symmetric contact matrix. The resulting contact rates can be interpreted as the average number of contacts per week per participant if the population were uniformly distributed over the age groups.

We compared contact characteristics between participants with and without frailty and between survey periods 1 and 2. For each contact, it was reported whether it was unprotected (e.g., without a face mask), whether it lasted longer than 15 minutes, whether it was closer than 1.5 metres, and whether the contact was physical. For each of these four high-risk types, the fraction of community contacts per participant was calculated. The differences between the four comparison groups were tested with a Mann-Whitney U test. In a similar way, the fraction of repeated community contacts per participant was analysed. Finally, the distribution over contact locations was compared visually between frailty status and the survey period.

## Results

### Study population

In total, 4914 invitations were sent out, and 820 questionnaires were returned, of which 730 were suitable for analysis. The 90 questionnaires not suitable for analysis were either empty with sometimes an explanation why the invitee was not able or willing to participate (64), without general information (7), without any contact data (7), or incomprehensible (12). Response rates (Suppl. S4) decreased with increasing age group and were higher for males than for females. The overall response rate for survey period 1 in April 2021 (17%) was higher than for survey period 2 in October 2021 (13%).

The two survey periods consisted of similar numbers of participants (Tab. 1). The age group distribution of the study population ranges from 17% for the 90+ age group to 23% for the 85-89 age group, close to the aim of 20% for each of the five age groups. More males (56%) than females participated. A large majority (95%) of the participants were born in the Netherlands. Based on the number of Dutch citizens with a first-generation migration background by age group [32], we would expect 53 participants with another country of birth in the study population. Instead, only 34 participants were not born in the Netherlands, which cannot be explained by stochastic effects, indicating a lower response rate in this group.

**Table 1:** Number of participants, stratified by survey round, age group, gender, country of birth, frailty, household size and education level, for all 730 participants.

| variable | value | n (%) |
| --- | --- | --- |
| Survey period | 1 (April 2021) | 345 (47.3) |
|  | 2 (October 2021) | 385 (52.7) |
| Participant age group | 70-74 | 142 (19.5) |
|  | 75-79 | 137 (18.8) |
|  | 80-84 | 155 (21.2) |
|  | 85-89 | 168 (23.0) |
|  | 90+ | 128 (17.5) |
| Participant gender | Female | 319 (43.7) |
|  | Male | 411 (56.3) |
| Country of birth | Netherlands | 694 (95.1) |
|  | Other country | 34 (4.7) |
| Frailty (GFI)<sup>a</sup> | 0-1 (non-frail) | 253 (34.7) |
|  | 2-3 (non-frail) | 218 (29.9) |
|  | 4-5 (frail) | 144 (19.7) |
|  | 6-7 (frail) | 69 (9.5) |
|  | 8+ (frail) | 42 (5.8) |
| Household size | 1 | 285 (39.0) |
|  | 2 | 424 (58.1) |
|  | 3+ | 8 (1.1) |
|  | Not applicable | 10 (1.4) |
| Education level | Primary or no education | 94 (12.9) |
|  | Secondary education (3 or 4 years) | 303 (41.5) |
|  | Secondary education (5 or 6 years) | 144 (19.7) |
|  | Higher education | 176 (24.1) |

Number (percentage) of participants with missing values for country of birth: 2 (0.3%), for frailty: 4 (0.5%), and for education level: 13 (1.8%)
<sup>a</sup>GFI: Groningen Frailty Indicator

Around one-third of the participants had a frail status, according to the GFI. Frailty was higher for females than for males in the same age group, and it increased with age but faster for males than for females (Fig. 1A). These findings agree with the literature (Suppl. S2) and results reported for the Tilburg Frailty Indicator of community-dwelling older adults [38], which is a frailty indicator similar to the GFI. Most participants (60%) lived with one or more household members. According to the number of Dutch older adults in long-term care facilities [33], we would expect 60 participants in care. Instead, only 10 participants indicated that they live in a nursing home or similar long-term care facility, emphasising that the study population is more representative of community-dwelling older adults. One-person households were more common for higher age groups and for females (Fig. 1B). The education level was, in general, higher for younger age groups and for males (Fig. 1C).

**Figure 1:**
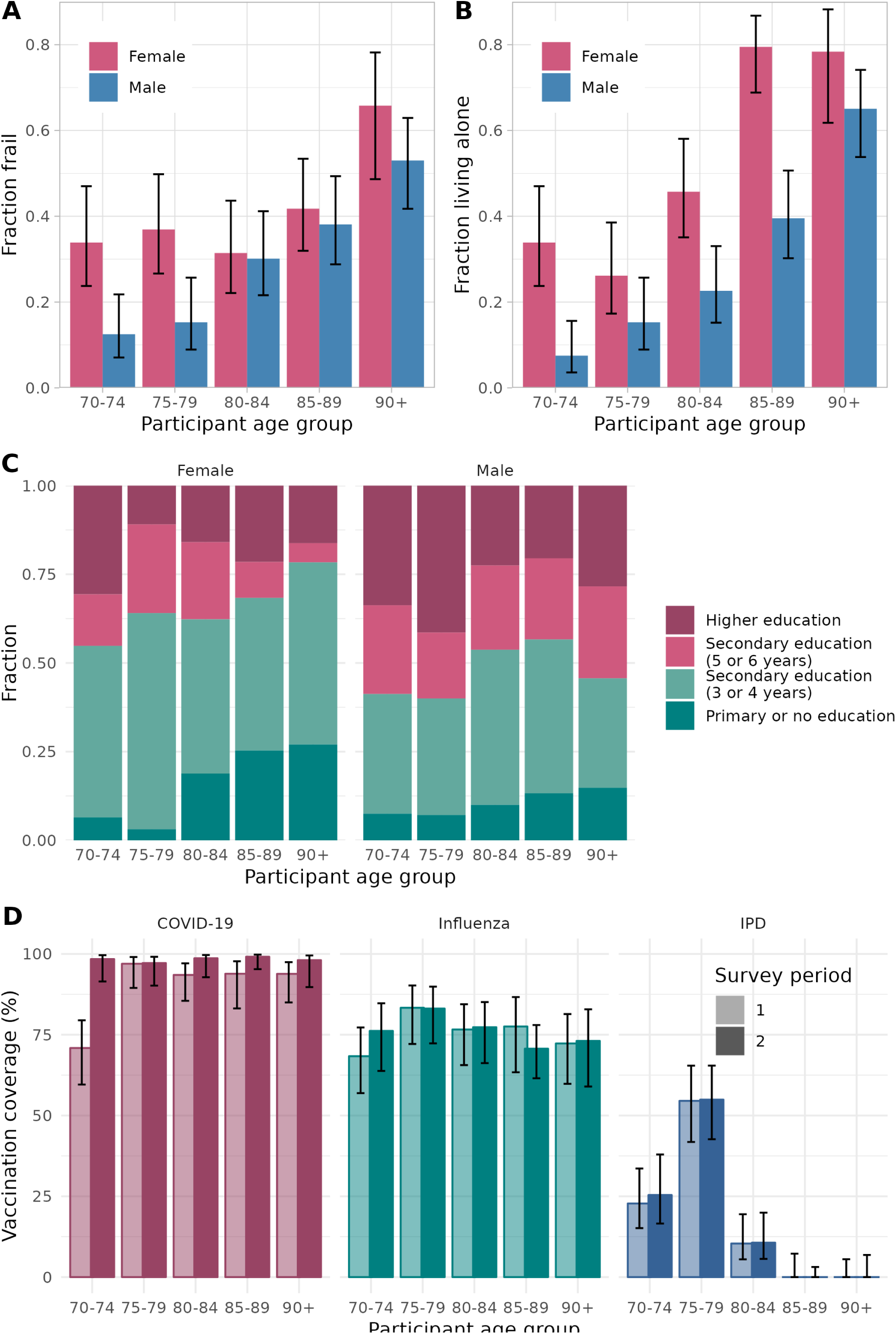
Participant characteristics of the study population consisting of 720 community-dwelling participants. (A) Fraction of participants with frailty by age group and gender. (B) Fraction of participants living alone by age group and gender. (C) Distribution of education level of participants by age group and gender. (D) Vaccination coverage against COVID-19, influenza and invasive pneumococcal disease (IPD) by age group and survey round.

Participants reported whether they had been vaccinated in the last 12 months against COVID-19, influenza, or IPD (Fig. 1D). The highest coverage was observed for COVID-19 in all age groups. Only the age group 70-74 fell slightly below other age groups in the first survey period in April 2021, as they had only just been invited for vaccination. The overall vaccination coverage in the study population in the second survey period in October 2021 (98%) was higher than the actual coverage reported for the 70+ population (93%) [34]. The overall vaccination coverage against influenza in the study population (75%) was higher than in the general 65+ population (73%) [35]. Nothing can be concluded from the IPD vaccination coverage because the IPD vaccination programme for older adults has only been implemented recently for specific age groups [36].

### Contact behaviour

For the contact behaviour analysis, 4 participants without frailty status, 10 participants in long-term care facilities, and 12 participants who participated for less than 5 days were excluded. Of the 704 included participants, 46 provided contact information for less than 7 days. The number of contacts on their missing days was imputed while taking reductions in the average number of contacts per day into account of 4% on each additional participation day and 20% on Sundays (Suppl. S3). The household size did not affect the number of community contacts per week and is not included in the full model. By comparing full and subset formulations, we found some evidence that the weekly number of community contacts of participants with and without frailty differed in survey period 1, though not statistically significant (p-value = 0.085, chi-square test) and differed significantly in survey period 2 (p-value = 0.023). The number of community contacts per week between the two survey periods differed significantly for persons with (p-value = 0.0050) and without (p-value < 0.0001) frailty. According to the full model, the expected weekly number of community contacts in survey period 1 was 14 (12 - 17) (mean and 95% confidence interval) for an average person with frailty and 19 (16 - 21) for an average person without frailty. In survey period 2, they were 21 (17 - 25) for an average person with frailty and 26 (23 - 30) for an average person without frailty.

When plotting the full model results with all covariates (Fig. 2), we noticed some remarkable patterns. The weekly number of community contacts in survey period 2 - with few COVID-19 measures - only slightly decreased with age in any stratum. Females without frailty had more contacts than females with frailty of any age, and males with frailty had the same number of contacts as a 12-year older male with frailty. In survey period 1, persons with frailty of all ages decreased their contacts to a similar extent compared to survey period 2, as demonstrated by the almost parallel lines in Fig. 2. For persons without frailty, however, the lines diverge, showing that the number of contacts in survey period 2 clearly decreased by age. This trend is obvious in both males and females.

**Figure 2:**
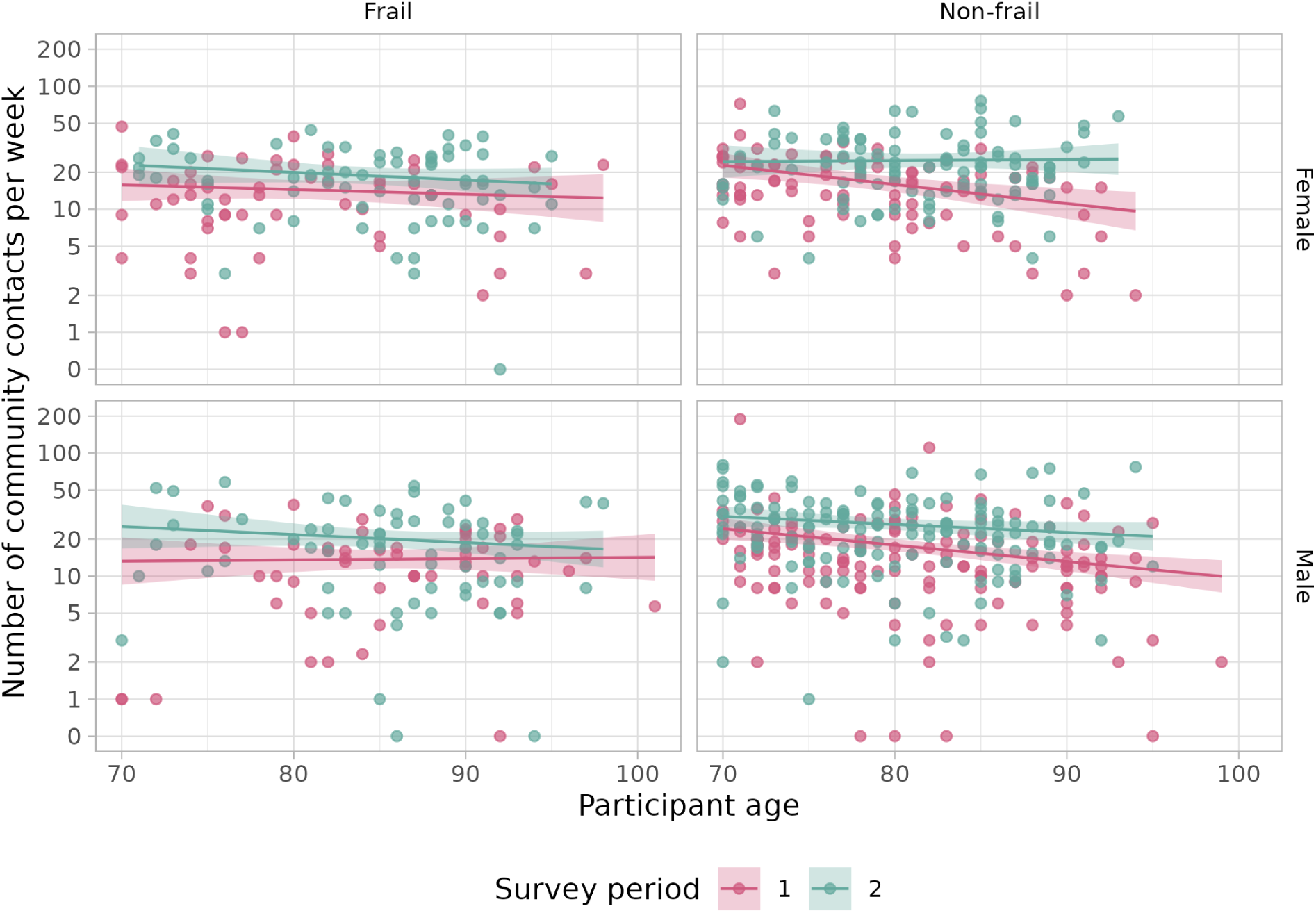
Weekly number of community contacts (i.e., persons contacted outside the household) per participant by age in survey periods 1 and 2. Plots show the data (one point for each participant) and model results (mean as solid line and 95% confidence interval as shaded area) by frailty (columns) and gender (rows).

The same effect is apparent in the contact matrices by frailty and survey period (Fig. 3). In all contact matrices, the highest contact rates are on the diagonal, meaning that participants interact mainly with people in the same age group. Participants with frailty had lower contact rates than participants without frailty, and in survey period 1, lower contact rates were observed than in survey period 2. The largest differences between survey periods 1 and 2 are seen for older participants without frailty. In survey period 2, they interacted mainly with other people of similar age, from which they refrained in survey period 1.

**Figure 3:**
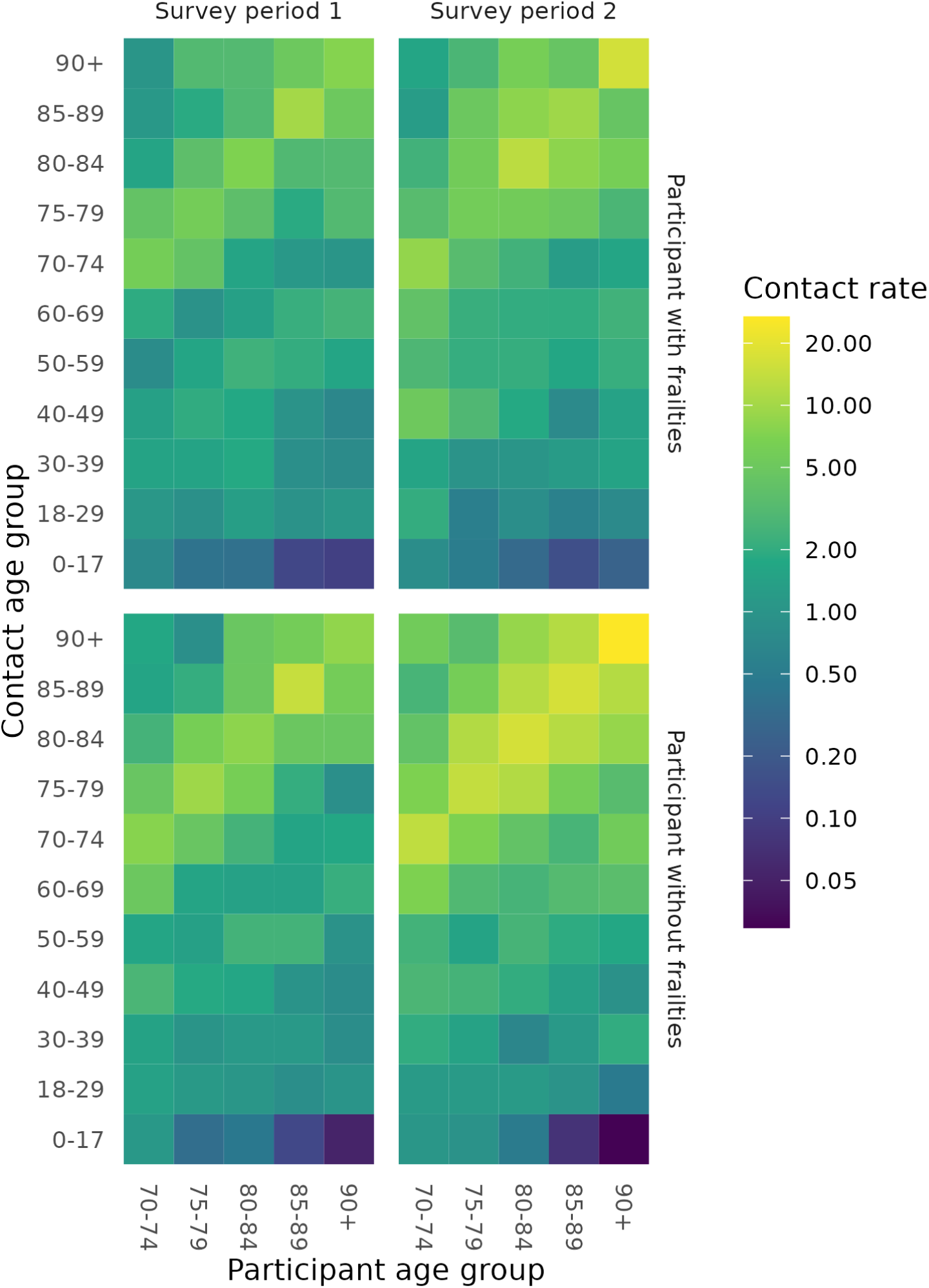
Contact matrices showing age mixing patterns for participants with and without frailty in survey periods 1 and 2. The contact rate can be interpreted as the average number of community contacts (i.e., persons contacted outside the household) per participant per week, if the population were uniformly distributed over the age groups.

### Contact characteristics

Contact behaviour change is not only reflected in the number of community contacts but also in the type of community contacts. For all types of contacts, participants shifted to more risky behaviour in survey period 2 compared to survey period 1: the fraction of community contacts per participant that were without protection, closer than 1.5 m, lasting over 15 min, and involved physical contacts increased significantly (Fig. 4). For the most part, differences between participants with and without frailty were not significant. One exception is that participants with frailty used protection such as face masks more often than participants without frailty in survey period 1. Another exception is that participants with frailty reported a significantly higher fraction of physical contacts than participants without frailty in both survey periods.

**Figure 4:**
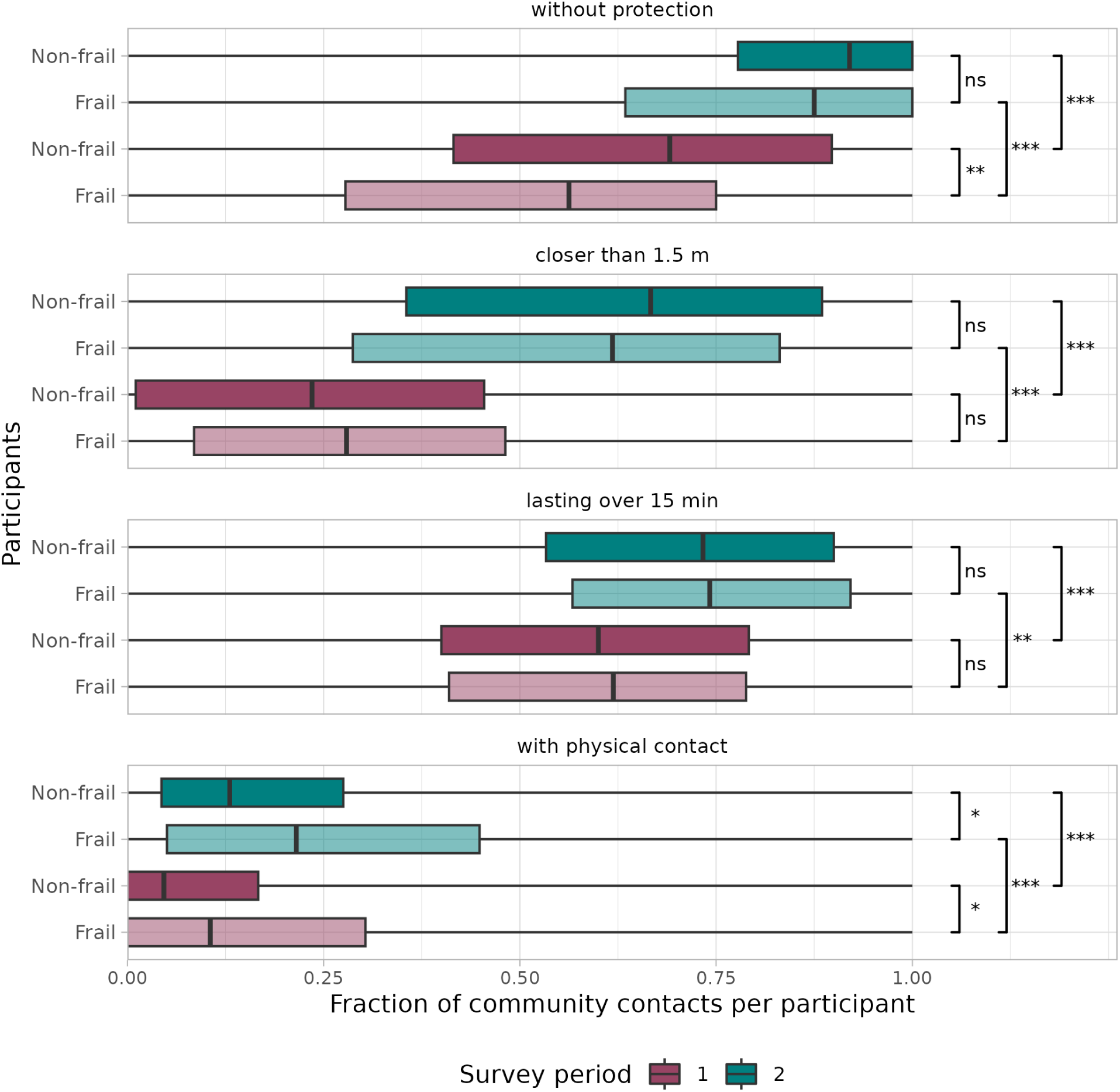
Fraction of community contacts (i.e., persons contacted outside the household) per participant for four risk behaviour factors: protection, distance, duration and physicality of the contact. Distinction is made between frailty of participants (transparency) and survey period (color). The whiskers of the boxplots extend to the minimum and maximum values. Significance levels are denoted by *** (p-value < 0.001), ** (p-value < 0.01), * (p-value < 0.05) and ns (not significant), according to the Mann-Whitney U test.

The location where contacts take place also differs by frailty status and survey period. Participants with frailty had a similar fraction of community contacts at home in both survey periods and always more than participants without frailty (Fig. 5). In survey period 2, participants without frailty had especially more contacts at leisure activities. The fraction of repeated community contacts, i.e., persons who are not members of the household that are contacted more than once a week, did not differ between persons with or without frailty. Persons who lived alone, however, did have relatively more repeated community contacts than persons who lived in a multi-person household (Suppl. S5).

**Figure 5:**
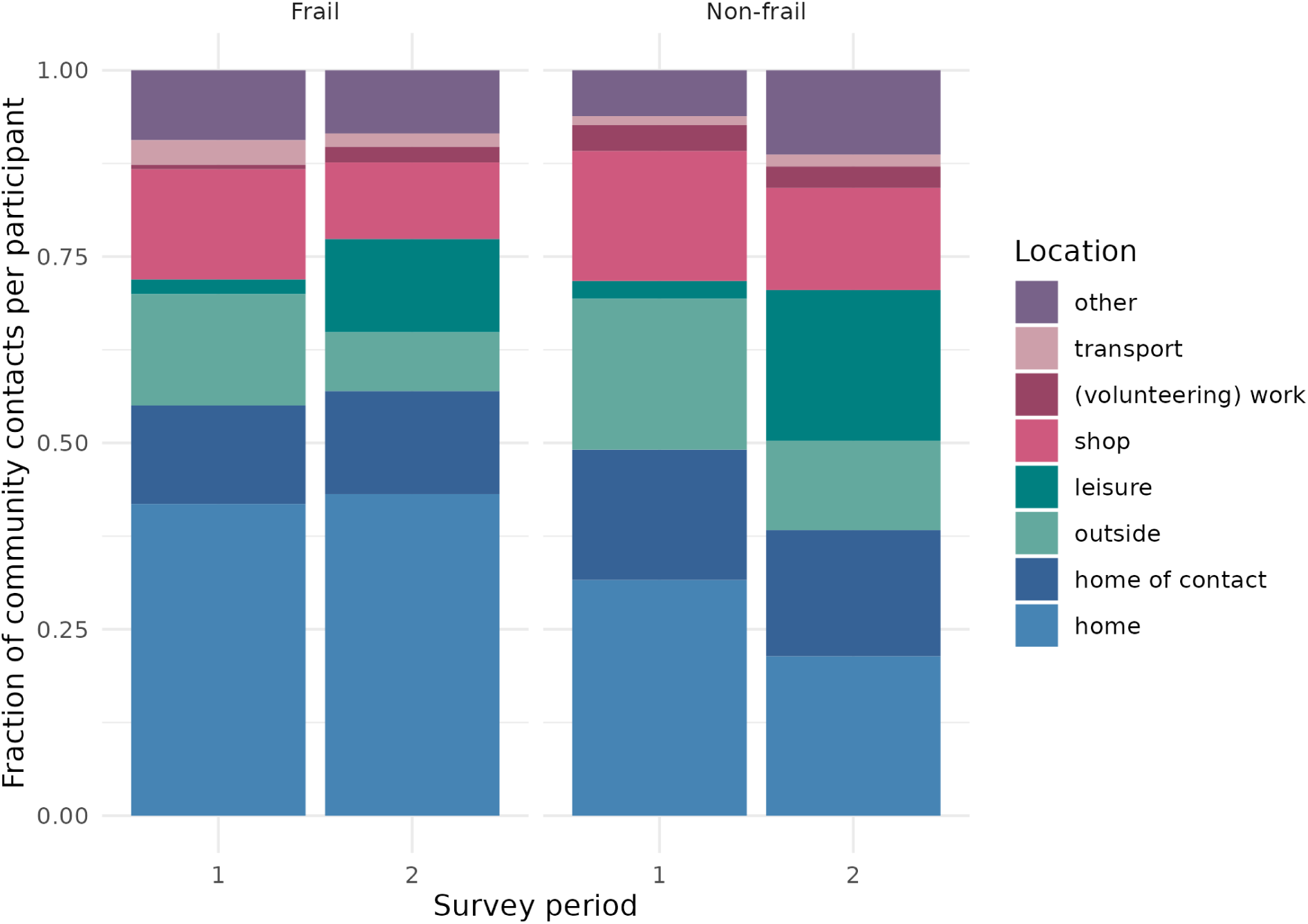
Fraction of community contacts (i.e., persons contacted outside the household) per participant by location for participants with and without frailty in survey periods 1 and 2.

## Discussion

In this survey, we studied how the contact behaviour of persons of 70 years and older in the Netherlands changed during periods with stringent and moderate COVID-19 measures in 2021 and how these changes differed for persons with and without frailty. We found that people with and without frailty had more community contacts in October 2021 with moderate measures than in April 2021 with stringent measures. During both survey periods, people without frailty had more community contacts than people with frailty. When we consider people with and without frailty separately, the number of community contacts only slightly decreased with age in the survey period with moderate measures. In this survey period, the observed frailty status of a participant is a more informative predictor of the number of contacts than age.

The two survey periods differed in both timing (spring vs. autumn) and in the intensity of COVID-19 measures (stringent vs. moderate). Of these two differences, we believe the latter is most essential in interpreting the observed results. Results of an earlier contact survey in 2016– 2017 [22] showed little effect of timing within the year on the number of contacts [3].

Therefore, it is unlikely that the differences found between the two survey periods are due to a seasonal effect, and the differences between survey periods are most probably caused by the difference in intensity of COVID-19 measures (stringent in April 2021 vs. moderate in October 2021) and the risk perception at the respective times of the surveys.

The results might have been affected by a poor representativeness of the study population. The study’s overall response rate of 15% was low. One reason for the low response rate is that the returned questionnaires were unmarked by design, which meant that it was not possible to send reminders to non-responders. Another reason is that participation required considerable effort to keep a contact diary for a full week, which made it difficult for invitees with cognitive issues or difficulties with reading and writing to participate. This may have led to a selection bias for participants without frailty. However, the frailty distribution by age and gender of the study population (Fig. 1A) agrees with results reported for the Tilburg Frailty Indicator of community-dwelling older adults [38], which leads us to believe that the study population is representative with respect to frailty of the community-dwelling older population.

While participants who were not born in the Netherlands were underrepresented in the study population as compared to the Dutch 70+ population, the distribution of frailty status and number of contacts for participants who were not born in the Netherlands did not differ from the other participants. The participants in long-term care facilities, who were excluded from the analysis, were more frail (8 out of 10 as compared 34% in the other participants) and had more contacts than the community-dwelling study population. We believe it unlikely that the observed differences in contacts between survey periods and between participant groups have been affected by unintended deviations from the representativeness of the 70+ community-dwelling study population.

The number of reported contacts per day by 70+ participants is within the order of magnitude of what would have been expected based on other surveys that address the general population. A possible exception is that in the second survey period, the 3.9 (3.7 - 4.0) contacts per day (mean and 95% bootstrap CI) is much lower than the 7.2 (6.2 - 8.5) and 7.0 (6.0 - 8.4) contacts per day found for 70+ participants in two previous studies in 2006-2007 [21] and 2016-2017 [22], respectively. Aside from the fact that the study populations and questionnaires were different, several other factors could explain this discrepancy. Most likely, this is because participants had not reverted back to pre-pandemic behaviour in October 2021. However, we cannot rule out other possible explanations: having to fill out the diary every day causes a fatigue effect (Suppl. S3), and correcting for this effect would increase the average number of contacts by 13%; due to COVID-19 information campaigns, participants better understand what constitutes a contact that could possibly lead to transmission, and they may have filled out the diary more conservatively compared to pre-pandemic contact surveys.

During both study periods, two other contact surveys were conducted among the general Dutch population. The PiCo survey was held three times per year among a representative sample of the population in the Netherlands [3,39]. The CoMix survey was held every two weeks on a selected internet panel [40,41]. The 70+ participants in the PiCo and CoMix surveys reported 2.3 (2.1 - 2.5) (mean and 95% bootstrap CI) and 2.5 (2.0 - 3.1) contacts per day around survey period 1 and 5.1 (4.7 - 5.5) and 4.8 (2.1 - 10.2) contacts per day around survey period 2. While these studies agree, they find a larger difference between the survey periods than this study: 2.8 (2.6 - 3.0) and 3.9 (3.7 - 4.0) contacts per day in survey periods 1 and 2, respectively. This could be an effect of differences in study setup, questionnaire, study population, or fatigue. For instance, the 70+ participants in both the PiCo and CoMix surveys are on average younger than in this study, where older age groups are oversampled. Summarising, where we find larger differences between the number of contacts as reported in the current study and the number reported in earlier studies, these can be explained by the different age composition of the surveys and by different contact behaviour in October 2021 as compared to pre-pandemic levels. None of these differences affect the interpretation of our findings.

In previous studies [26,27], it was observed that the number of contacts of older adults decreases with age. The results of this study show that in survey period 2, which does not fully but most closely resemble a normal situation, the number of community contacts only slightly decreased with age (Fig. 2). Because persons with frailty have on average fewer contacts than persons without frailty, the decrease by age as observed in previous studies without frailty distinction could also be explained by persons transitioning from non-frail to frail status as they grow older. Especially for females, frailty is a better predictor of the number of contacts than age. In survey period 1, with more stringent COVID-19 measures, the number of contacts of persons without frailty did decrease with age. These older persons without frailty interact mainly with persons of similar age (Fig. 3) who are increasingly frail (Fig. 1A). As a consequence, their behaviour change indirectly protected their peers with frailty. Younger persons without frailty did not decrease their contacts as much, possibly because they did not interact with persons with frailty as much, and persons with frailty themselves all decreased their contacts, probably to protect themselves.

The results of this study reveal how social distancing measures affected the contact behaviour of persons with and without frailty aged 70 years and older during the COVID-19 pandemic in the Netherlands, with the largest decrease in number of contacts for the oldest participants without frailty. These results can be useful in different ways. Frailty questions could be included in contact surveys for the general population, as frailty is an additional indicator for the number of contacts. Stratifying infectious disease models by frailty could increase knowledge on how people with and without frailty are affected under different control scenarios, although it would first be necessary to know how people with and without frailty mix. These results can also be instrumental in public health policy, for instance, by shaping information campaigns on social distancing measures. As the population in many countries ages rapidly, it is becoming ever more important to take the frailty differences of older adults into account to be prepared for future pandemics.

## Declarations

### Ethics approval and consent to participate

The Medical Research Ethics Committee (MREC) NedMec confirmed that the Medical Research Involving Human Subjects Act (WMO) does not apply to the SCONE (Studying Contacts of Elderly) study (research protocol number 23-051/DB). Therefore, official approval of this study by the MREC NedMec is not required under the WMO.

The contact survey was performed in accordance with relevant guidelines and regulations. Participants were informed that by returning the original questionnaire to RIVM, they were consenting to participate in the study. The initial processing of personal data by RIVM is lawful since it is necessary for the performance of a task carried out in the public interest (Article 6, paragraph 1, under e, of the GDPR).

### Consent for publication

The participants were informed that the data and results would be made publicly available in an anonymous way, so that individual participants could not be (re)identified. In the context of making the data and results publicly available for scientific or statistical purposes, the principle of data minimization is being respected. Whenever possible, the data is minimised by aggregating open, easily identifiable, and unnecessary records (Article 5, Paragraph 1, under b, and Article 89, Paragraph 1, of the GDPR).

### Availability of data and materials

The cleaned data is published online at https://doi.org/10.5281/zenodo.7649375 [30]. All code for data cleaning and analysis is publicly available at https://github.com/rivm-syso/SCONE [31].

### Authors’ contributions

J.B, F.E.F., N.H. and J.W. designed the study, J.B. collected the data, J.B. and J.v.d.K. did the statistical analyses, J.B. wrote the main manuscript text. All authors reviewed the manuscript.

## Acknowledgments

The authors would like to thank all participants of the SCONE (Studying Contacts of Elderly) study for their invaluable input, Inge Besemer for processing the data, and Kylie Ainslie and Brechje de Gier for critically reading the manuscript.

