## Supplementary material for "Contact patterns of older adults with and without frailty in the Netherlands during the COVID-19 pandemic"

S1 SCONE study design

S2 Frailty determinants

S3 Fatigue and day-of-week effect

S4 Response rates

S5 Repeated contacts

### S1 SCONE study design

The aim of the SCONE (Studying CONtacts of Elderly) survey is to learn about contact behaviour of persons of 70 years and older, by means of keeping a contact diary for a full week. The study design consists of questionnaire development, participant recruitment, and data entry, cleaning and publication.

#### Questionnaire development

To keep a diary for a full week, prospective participants were sent a 24-page booklet, consisting of an extensive explanation and the questionnaire itself (Suppl S5). The explanatory part starts with the motivation for the study and the request to participate in it. A manual how to fill out the questionnaire explains all questions one by one. Next, an example situation is described with the correct way to enter this in the diary. Some examples are given on what constitutes a contact, e.g. short face-to-face conversations or physical contacts, and what does not, e.g. telephone calls or one-word greetings to the neighbor. The explanation finishes with a list of frequently asked questions, mainly to reiterate the directions of the previous pages.

The questionnaire to be filled out consists of a general part and two opposite pages for each day of the week. The general questions collect the variables:

- Year of birth
- Age (as check for year of birth)
- Gender
- Country of birth
- Education level (4 options, tailored to school types common in 1940’s, 1950’s and 1960’s)
- Household size (with option to tick “not applicable” for persons living in nursing home)
- Vaccination status against COVID-19, influenza and pneumococcal disease (for vaccines received in last 12 months)
- 15 questions of Groningen Frailty Indicator [1] to determine the frailty status

For each day of the week, the participant fills out the date and indicated whether they had any contacts on that day. The possibility to mark ‘no contacts’ is essential to distinguish participants without any contacts from participants that did not fill out the questionnaire on that day. The participants with contacts report for each unique person they contacted that day:

- Name or description (to identify persons who are also contacted on other days)
- Age (either exactly, or an estimated age range)
- Gender
- Household member: yes/no
- Physical contact: yes/no
- Protected contact (e.g. by face mask or transparent shields): yes/no
- Contact proximity: more or less than 1.5 meters

- Contact duration: longer or shorter than 15 minutes
- Contact location: at home, at other person's home, at (volunteering) work, in transit, during leisure time, in a shop, outside, other (combinations allowed); space to specify other location

Per day a maximum of 21 contacted persons could be filled out, using one row per contact. A small space to describe more contacts was provided at the bottom, but in practice many participants used one row to describe groups of persons they encountered during a specific activity (shopping or at the bridge club).

For ease of filling out, no tick boxes were used in the yes/no questions, but "yes / no" was printed, and the applicable option should be circled. Each day of the week had a different light background color to distinguish them.

The first version of the questionnaire was tested in small panel of 5 volunteers (3 females, 2 males, ages ranging from 71 to 78). This led to some modifications, mainly in clarifications of the definition of a contact. Also, the list of examples of what constitutes a contact and what does not, was added.

### Participant recruitment

To invite persons of 70 years and older to participate, they were randomly selected from the Personal Records Database [2] that contains the personal data of people who live in the Netherlands. The selection occurred according to a prespecified distribution over gender and 5 age groups (70-74, 75-79, 80-84, 85-89, 90+), aiming to have at least 50 participants in each stratum. For the second round of the study, this distribution was adjusted according to the observed response rates in the first round. Participants were never invited for both rounds, to ensure independent cross-sectional data.

All selected persons received an invitation, except when they were deceased at the time of selection. The invitees receive a letter that explains the purpose and conditions of the study, the booklet containing the questionnaire and an envelope to return the filled-out questionnaire. A telephone number and email address were provided to get more information.

In the invitation it is emphasized that participation is voluntary. To minimize the collection of personal data, the questionnaires are unmarked, so it is not known which invitees participate. The obvious but accepted drawback of this set-up is that it is not possible to send a reminder, which will decrease the achievable response rate. Participants were informed that by returning the original questionnaire to the RIVM, they were consenting to participate in the study. They were also informed that the data and results would be made publicly available in an anonymous way, so that individual participants could not be (re)identified.

### Data entry

All return envelopes that were received would get a participant id, also when containing an empty questionnaire or a letter of non-participation. Questionnaires with at least the age and gender of the participant and at least one day with comprehensible contact data, were deemed suitable for analysis. The data of these suitable questionnaires were entered in a Microsoft Access database.

The database contained simple checks during data entry. For example it checked whether age and year of birth agree, and whether the participant's age is 70 years or more. It was checked whether all data fields have a value. When a participant skipped a question, this should be indicated during data entry (selecting the option 'empty'). Without this requirement, it is impossible to know if an empty field is due to the participant skipping the question or the data entry clerk forgetting to enter the data.

The names of contacted persons were not a part of the data to be entered. Instead, only persons who were contacted more than once in a week got a unique letter to identify them as repeated contacts. At the end of a data entry day, more checks were run in an R script. For example it checked whether a repeated contact had the same age and gender on each reported day, whether no duplicate participant id's were entered, and whether a repeated contact occurred at least twice. The script produced a report with errors, that could be corrected the next day before starting new data entry.

Data entry into the Access database was performed by one person. Another person carried out a data quality check using the method of double data entry [3]. The entered data was checked for each contact diary until ten subsequent diaries were found without errors, after which only every tenth contact diary was checked. If an error was discovered, the sequence of finding ten subsequent errorless contact diaries was started again.

### Data cleaning and publication

The entered data is reorganized to comply with the general format used in studies that are published on socialcontactdata.org. Because of the longitudinal data for both participants (filling out the diary daily during a week) and contacts (repeated contacts during the week), an alternative convention is adopted to be able to identify unique participants and contact persons. A typical participant id consists of one digit for the study round (1 or 2), and 3 digits for the participant. A typical contact id consists of one digit for the study round (1 or 2), 3 digits for the participant, and 3 digits for the contact person. To distinguish the different days of the week, additional id variables concatenate the participant id and contact id with 1 digit for the day of the week (1 to 7) when the participant filled out the diary, or when the contact occurred.

The GFI score is calculated for each participant, provided they have filled out at least 13 of the 15 GFI questions. The GFI score ranges from 0 to 15, and participants with a GFI score of 4 or higher are regarded as frail [1].

The data was checked for inconsistencies. Two such inconsistencies were corrected. When participants reported they lived alone or in a nursing home, contacts can never be a household member (corrected in 144 instances). When a participant does not know the exact age of a household member, this person is likely not a household member (corrected in 62 instances). Other inconsistencies were checked but not corrected. Not all household members were reported every day the participant filled out the diary (16 household members, 9 of which were only missed one day). Some participants mention household members that don’t agree with household size (99 participants, of which 95 report a two-person household without mentioning a spouse as household member) The data cleaning script can be found on GitHub [4], and the cleaned data is published on Zenodo [5] following the data format of socialcontactdata.org. The paper diaries are destroyed after the end of the project.

### S2 Frailty determinants

As the aim is to study contact behaviour of people with and without frailty, it is important to understand which participant characteristics determine frailty. The participants that live in a nursing home or a similar care facility are excluded to focus this analysis on community-dwelling older adults. Odds ratios are calculated using multiple logistic regression, in pseudo-notation:

$$\text{frailty} \sim \text{age} * \text{gender} + \text{bornNL} + \text{education} + \text{household} + \text{period} \qquad (1)$$

with frailty (0 or 1), participant age (numeric), participant gender (Male or Female), born in the Netherlands (yes or no), education level (4 levels), household size (1 or 2+) and survey period (1 or 2). Because of the interaction term between age and gender, results for age are given separately for males and females, and results for gender are given at the mean age of the study population.

After excluding 4 participants without frailty status, we used multiple logistic regression (Eq. (1)) to study which participant characteristics are associated with frailty. Frailty increased with age, but the rate was higher for males than for females (Fig. 1). In general, females had a higher frailty than males of the same age. Persons living alone had a higher frailty than persons with household members, although this difference was not statistically significant. Persons with at least a secondary education had a lower frailty than persons with no or only primary education, but this effect was only significant when compared with persons who had attended 5 or 6 years of secondary education. There was no significant effect of country of birth or survey period on frailty.

Previous studies have found that frailty is higher for older persons, for females compared to males of the same age, and for persons living alone [1,6–8]. In our analysis of frailty determinants these effects were confirmed for age and gender. The survey period did not influence frailty, even though the level of COVID-19 measures differed between the survey periods. During the first COVID-19 wave in 2020, a higher frailty is reported mainly in the social domain [9], but this effect may have subsided during survey period 1 in April 2021 when COVID-19 was perhaps perceived as a lower risk.

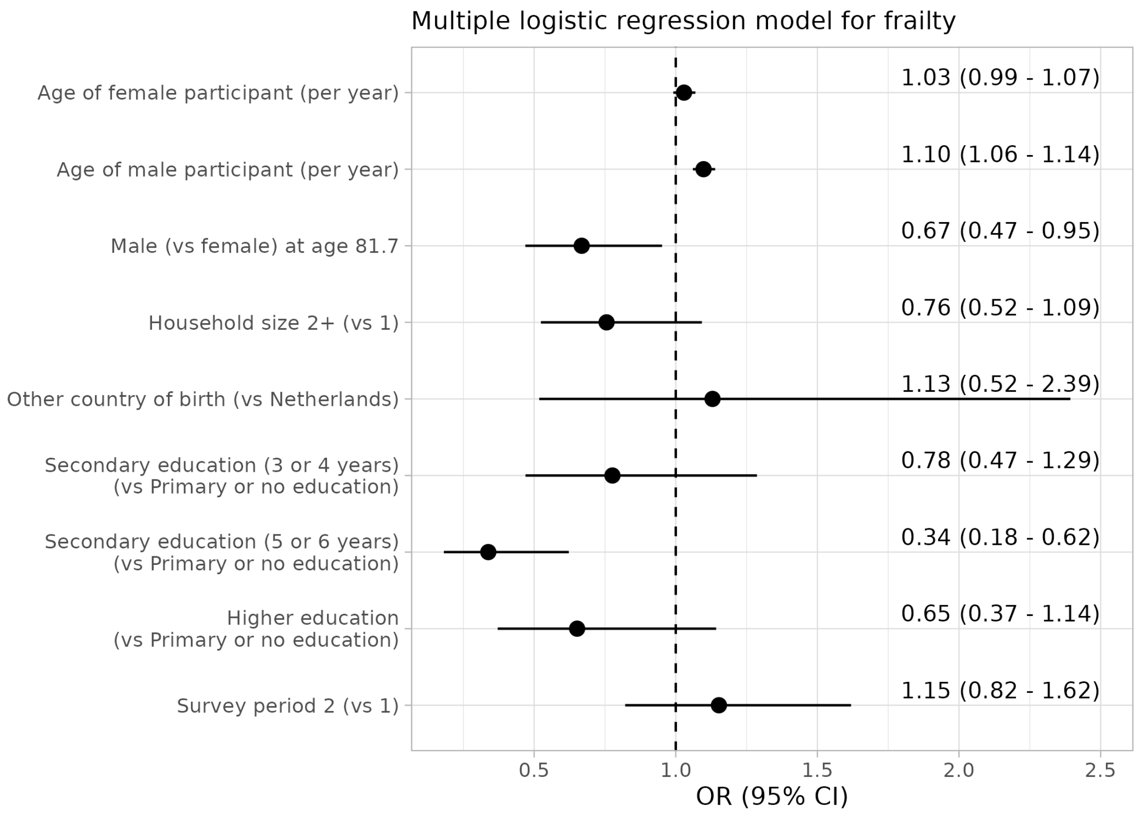


Figure 1: Forest plot of odds ratios (OR) and 95% interval for frailty determinants

### S3 Fatigue and day-of-week effect

In previous contact surveys [10,11] a fatigue effect was observed, where participants tend to report less contacts the longer they participate. In our study, a similar fatigue effect could play a role over the course of the participation week, but differences in reported numbers of contacts could also be caused by a weekday effect. An additional complication is that most participants start on a Monday, as this was the first page of the contact diary (Fig. 2).

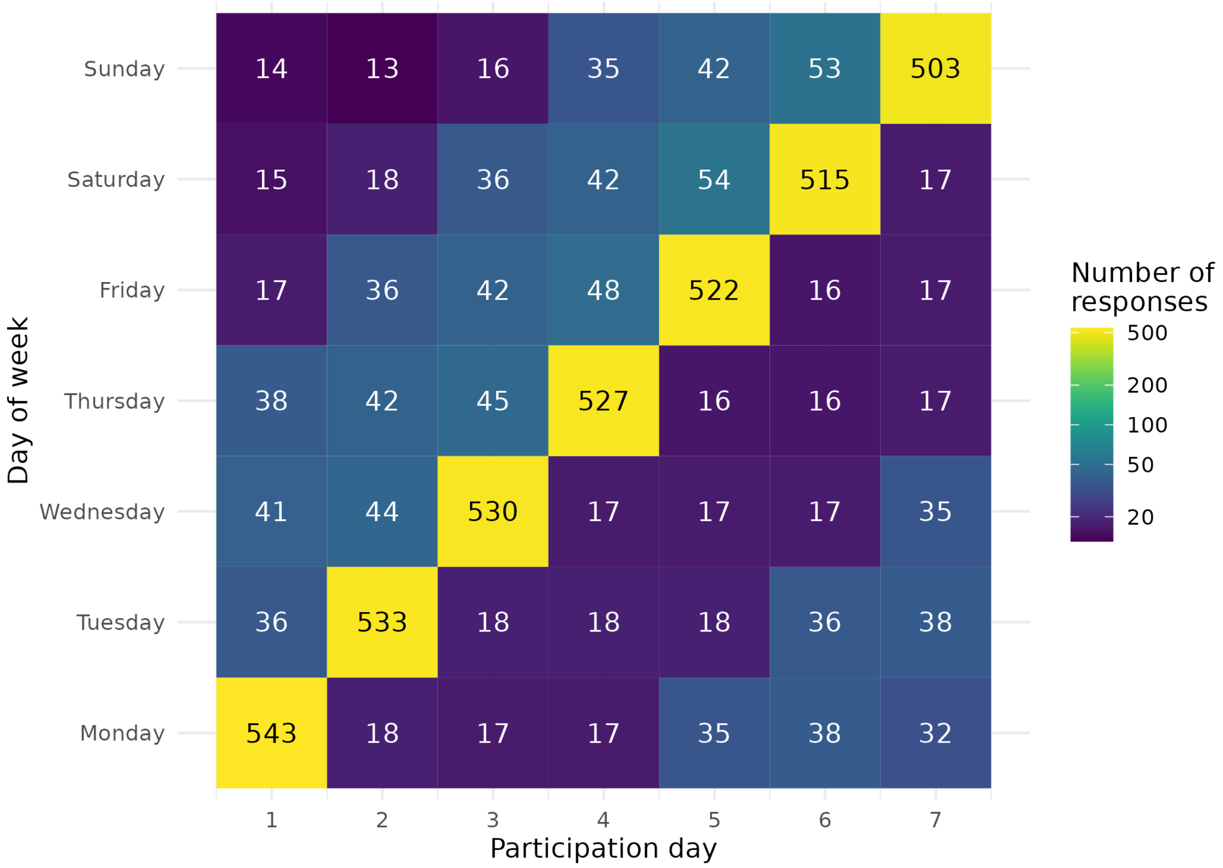


Figure 2: Number of responses by day of the week and participation day, for participants that participated at least 5 days. The majority of participants (77%) start the survey on a Monday.

To disentangle the fatigue and day-of-week effect, the number of daily community contacts $n_{\text{contacts}}$ is modelled by a generalised linear mixed model, where the number of contacts is assumed to follow a negative binomial distribution, in pseudo-notation:

$$n_{\text{contacts}} \sim \text{participationday} + \text{dayofweek} + b_{\text{id}}$$

with participationday (integer days), dayofweek (categorical) and $b_{\text{id}}$ a random effect for each participant that absorbs the effects of age, gender, frailty and survey period. The results show that the fatigue effect is around 4% per participation day (Fig. 3). For a full week of participation, this would mean that the actual number of contacts was 13% higher than the observed number of contacts. Furthermore, the numbers of contacts are similar for most days of the week, but on Sundays significantly fewer community contacts are reported.

Of the 704 participants included in the analysis, 46 participants participated 5 or 6 days. For these participants the number of community contacts on the missing days is imputed with the fitted model.

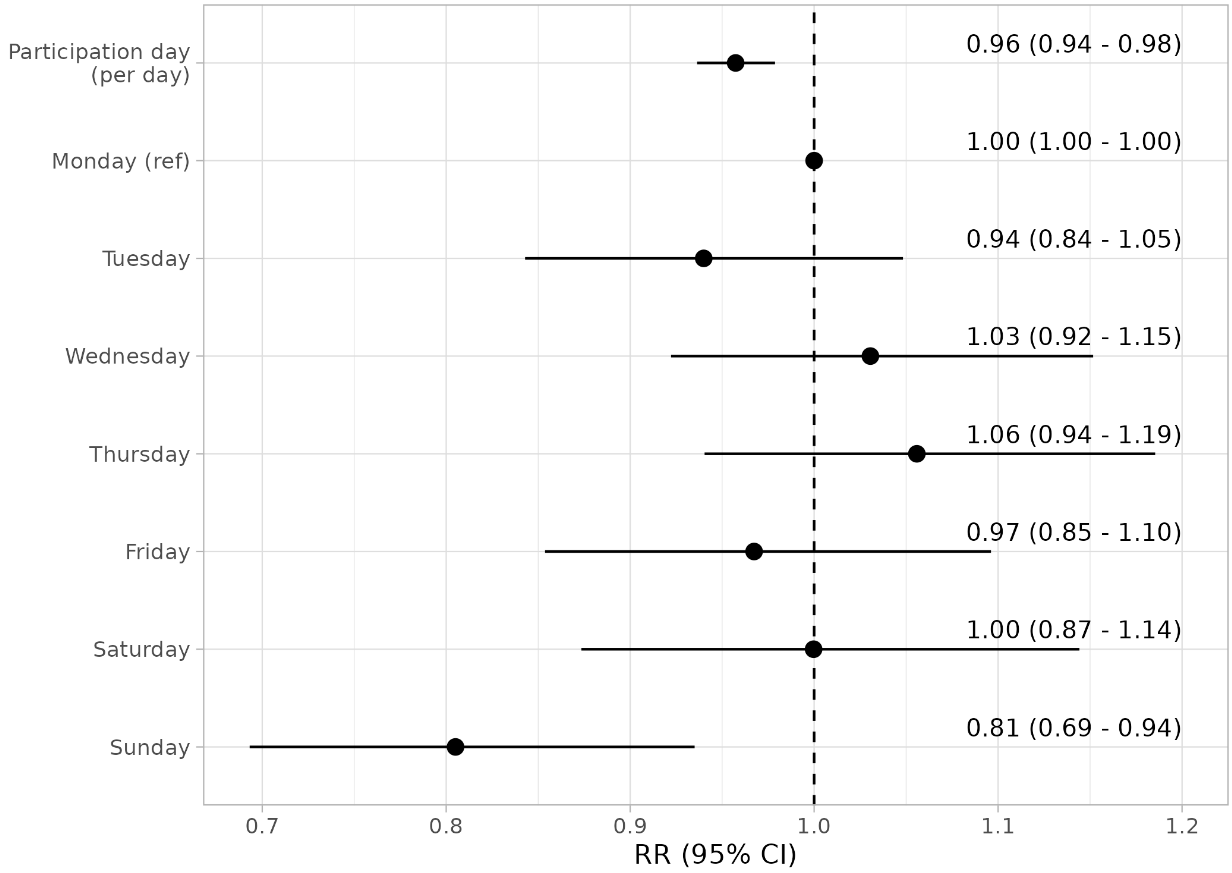


Figure 3: Forest plot of rate ratios (RR) and 95% interval for effect on the daily mean number of contacts by the day of the week and participation day (i.e. fatigue).

### S4 Response rates

| Survey period | Age group | Female | | | Male | | | Total | | |
|---|---|---|---|---|---|---|---|---|---|---|
| | | invited | included | % | invited | included | % | invited | included | % |
| 1 | 70-74 | 125 | 43 | 34 | 150 | 36 | 24 | 275 | 79 | 29 |
| | 75-79 | 150 | 34 | 23 | 169 | 32 | 19 | 319 | 66 | 21 |
| | 80-84 | 170 | 30 | 18 | 199 | 47 | 24 | 369 | 77 | 21 |
| | 85-89 | 197 | 22 | 11 | 246 | 29 | 12 | 443 | 51 | 12 |
| | 90+ | 241 | 17 | 7 | 328 | 55 | 17 | 569 | 72 | 13 |
| | **total** | **883** | **146** | **17** | **1,092** | **199** | **18** | **1,975** | **345** | **17** |
| 2 | 70-74 | 149 | 19 | 13 | 200 | 44 | 22 | 349 | 63 | 18 |
| | 75-79 | 249 | 31 | 12 | 248 | 40 | 16 | 497 | 71 | 14 |
| | 80-84 | 296 | 41 | 14 | 245 | 37 | 15 | 541 | 78 | 14 |
| | 85-89 | 444 | 59 | 13 | 441 | 58 | 13 | 885 | 117 | 13 |
| | 90+ | 384 | 23 | 6 | 283 | 33 | 12 | 667 | 56 | 8 |
| | **total** | **1,522** | **173** | **11** | **1,417** | **212** | **15** | **2,939** | **385** | **13** |
| **total** | **total** | **2,405** | **319** | **13** | **2,509** | **411** | **16** | **4,914** | **730** | **15** |

### S5 Repeated contacts

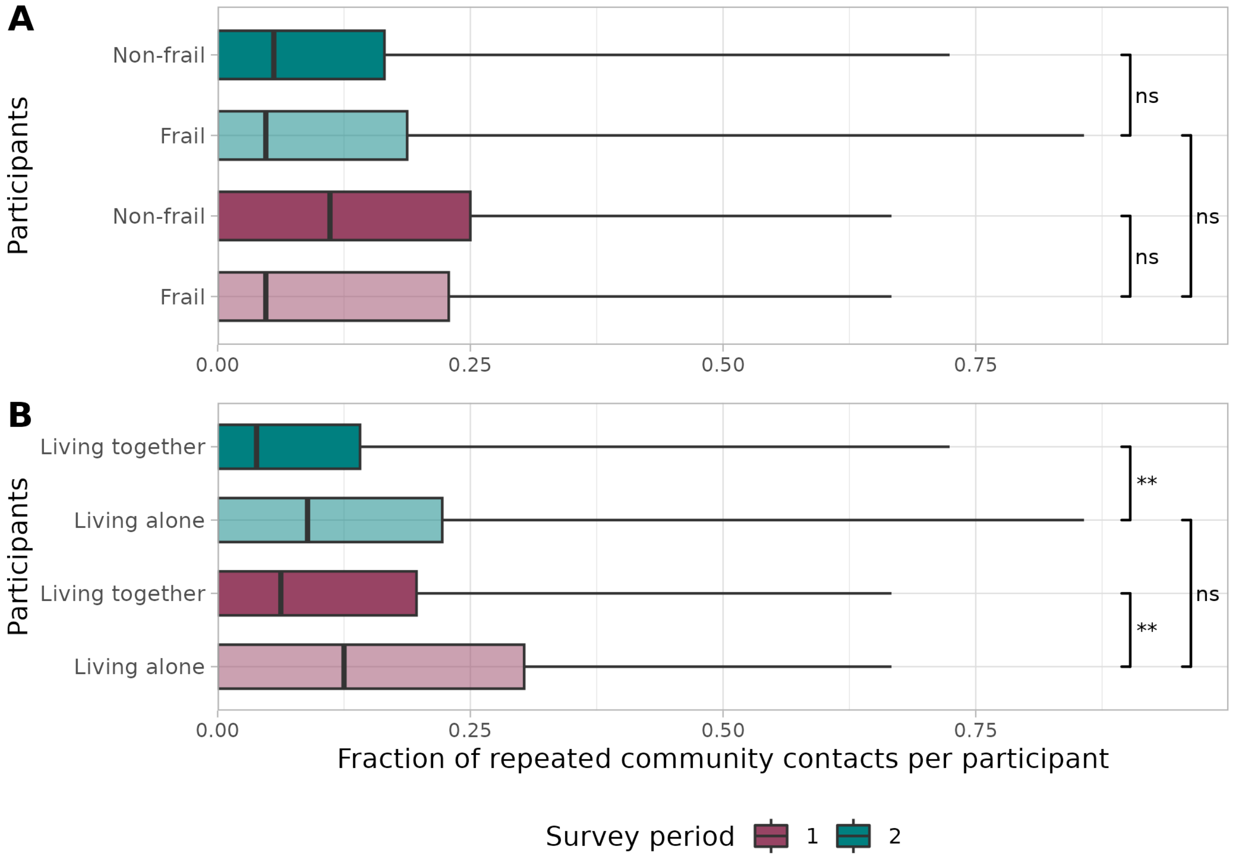


Figure 4: Fraction of repeated community contacts per participant by survey period (color) and frailty status (top) or living situation (bottom). The whiskers of the boxplots extend to the minimum and maximum values. Significance levels are denoted by *** (p-value < 0.001), ** (p-value < 0.01), * (p-value < 0.05), and ns (not significant), according to the Mann-Whitney U test.
